# Analysis of the effects of statin therapy on clonal dynamics in clonal haematopoiesis of indeterminate potential: insights from the English Longitudinal Study of Ageing

**DOI:** 10.1101/2025.04.15.25325668

**Authors:** Ellen Nuttall Musson, Yvette Hoade, Phoebe Dace, Javier Herrero, Spiros Denaxas, Andrew Steptoe, Elspeth Payne

**Affiliations:** Department of Haematology, UCL Cancer Institute, University College London, London, UK; The Genome Function Laboratory, The Francis Crick Institute, London, UK; Bill Lyons Informatic Centre, UCL Cancer Institute, University College London, London, UK; Institute of Health Informatics, University College London, London, UK; British Heart Foundation (BHF) Data Science Centre, Health Data Research UK, London, UK; NIHR Biomedical Research Centre at University College London Hospitals (UCLH) NHS Foundation Trust, London, UK; Department of Behavioural Science and Health, Institute of Epidemiology and Health Care, University College London, London, UK

## Abstract

Clonal haematopoiesis of indeterminate potential (CHIP) is the acquisition of somatic mutations in leukaemia-associated genes in haematopoietic progenitors with age. It increases the risk of haematological malignancy (HM), cardiovascular disease (CVD) and mortality mediated by CHIP-associated inflammation, with larger clones posing higher risks. Statins have been found to reduce the risk of progression from myelodysplastic syndrome to acute myeloid leukaemia and have also shown efficacy in vitro against TET2 deficient AML cell lines. However, their effect on CHIP has not been described. This study characterises the English Longitudinal Study of Ageing as a novel longitudinal CHIP cohort, through genetic analysis of 13270 longitudinal peripheral blood samples from participants aged over 50. Using logistic and robust regression analysis, we show that statin therapy is associated with reduced TET2 CHIP clonal expansion in a gene specific manner. We also find that statin primary prevention is associated with significantly lower incidence of myocardial infarction and stroke in individuals with CHIP compared to controls. These findings provide evidence that a commonly prescribed mediation with a well characterised safety profile may modify the natural history of TET2 CHIP, thereby mitigating its associated health risks.

---

Age-related clonal haematopoiesis of indeterminate potential (CHIP) is a well described pre-malignant phenomenon that increases the risk of heamatological malignancy (HM), cardiovascular disease (CVD) and all-cause mortality, with larger clones (variant allele frequency (VAF) > 10%) conferring higher risk of adverse health outcomes(1–3).

Research on intrinsic clonal determinants of CHIP clonal growth have indicated that gene involvement is important, with DNMT3A clones expanding relatively slowly, *TET2* clones expanding at a rate of approximately 10% per year and clones carrying somatic mutations in splicing factor genes expanding more rapidly(4). Multiparameter models based on mutational data and a broad range haematological and biochemical laboratory parameters have been constructed to predict risk of myeloid neoplasm(5). However, little has been published on extrinsic determinants of CHIP clonal growth, such as commonly prescribed medications.

Statins have been found to modify progression of myeloid disorders in large epidemiological studies, with statin therapy associated with a reduced risk of progression from myelodysplastic syndrome (MDS) to acute myeloid leukaemia (AML)(6). Further, statins have been shown to promote apoptosis in *TET2* deficient leukaemia cells in vitro, via compounded inhibition of the cholesterol synthesis pathway(8). A recent study also suggested that dyslipidaemia is associated with incident *TET2* CHIP, although importantly the association of statin therapy itself was not included(7). We therefore sought to understand how statin therapy affects CHIP clonal dynamics in the English Longitudinal Study of Ageing (ELSA) as a novel CHIP cohort.

ELSA is a longitudinal cohort that has been running since the year 2002. Individuals aged over 50 are invited for follow up at two-yearly health questionnaires(9) and are invited for peripheral blood sampling every 4 years. Cohort demographics from the first blood-sampling timepoint (ELSA wave 2) of 6231 individuals are shown in Figure 1A.

**Figure 1.**
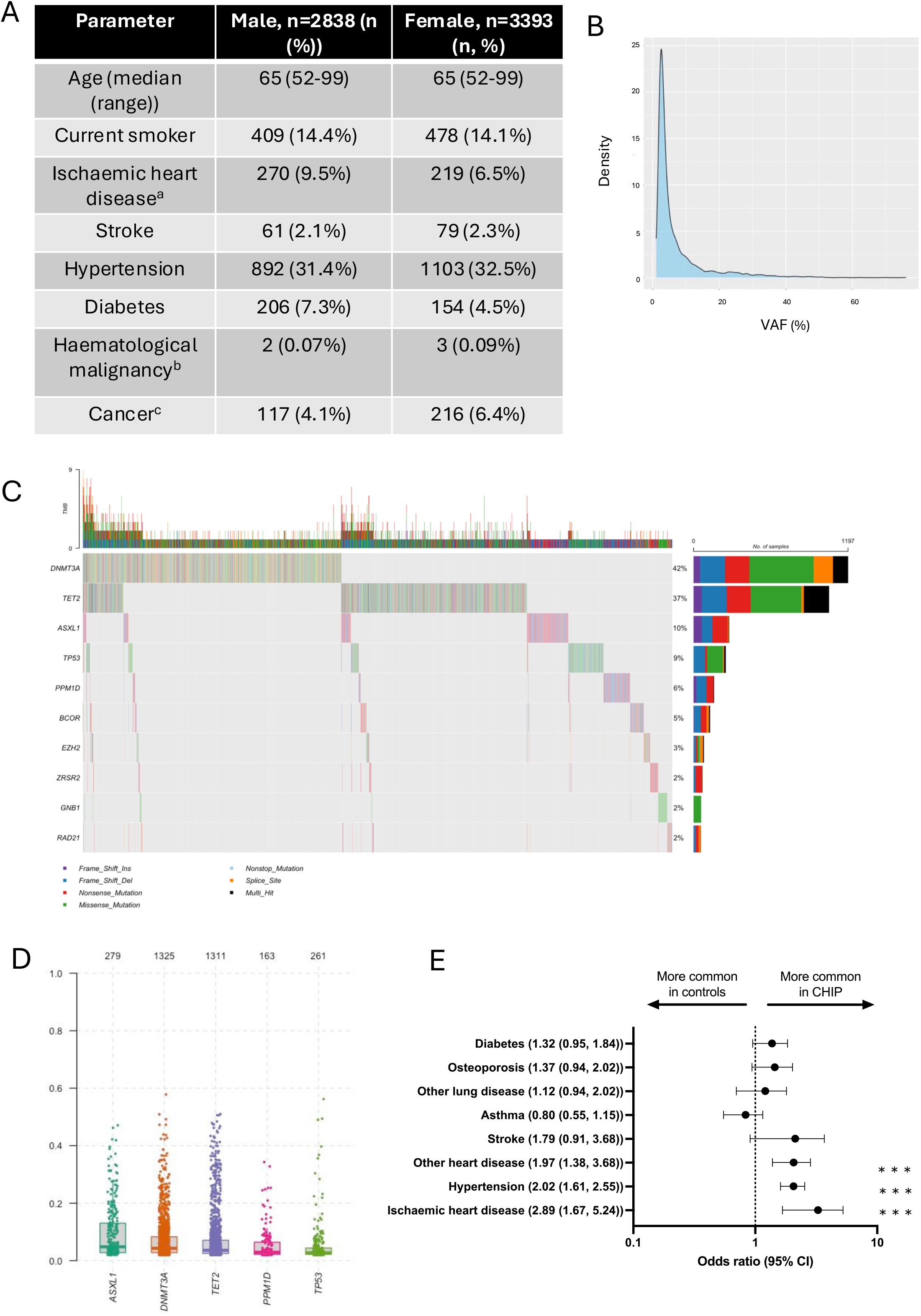
Characterisation of ELSA as a longitudinal CHIP cohort. A) ELSA demographics in wave 2 (2004, the first wave of peripheral blood sampling) by sex. ^a^Ischaemic heart disease is defined as a history of angina or myocardial infarction. ^b^Haematological malignancy is defined as a self reported diagnosis of leukaemia or lymphoma. ^c^Cancer comorbidity is defined as a history of any cancer in the last 2 years. B) Density plot of observed VAFs across the whole ELSA cohort. C) Oncoplot detailing the mutational profile of the ELSA cohort (including mutation count, type, co-occurrence and frequency) of the ELSA cohort in the 10 most commonly mutated genes. D) VAFdistributions of the five most commonly mutated genes in the ELSA cohort. E) Odds ratios and 95% confidence intervals (CI) for common comorbidities in CHIP cases and controls, matched for age, sex and smoking status. Ischaemic heart disease is defined as a history of angina or myocardial infarction, non-ischaemic heart disease define as arrhythmias and valvular heart disease, other lung disease defined as self reported non-asthma lung disease. *** p < 0.001.

A total of 13270 peripheral blood samples were screened for the presence of CHIP using a custom myeloid amplicon panel (Table S1) covering genes previously implicated in CHIP and myeloid malignancy. The mean coverage per sample across all amplicons was 1395. Methods of variant calling are provided in the supplementary material. Twenty-one percent (2846 samples) had detectable CHIP mutations at a VAF >=2%, with 3989 variants detected (Table S2). The mutational profile of the ELSA cohort is shown in Figures 1 B-D. In keeping with other published cohorts, the most commonly mutated genes were *DNMT3A* and *TET2* (accounting for 42% and 37% of the mutations detected, respectively (Figure 1C), with the majority of VAFs<10% (Figure 1B) (1,4).

To understand the epidemiological associations of CHIP in ELSA, CHIP cases and controls were matched on age, sex and smoking status and odds ratios for common comorbidities were calculated Figure 1E. CHIP was associated with an increased risk of ischaemic heart disease, non-ischaemic heart disease and also hypertension, indicating that the ELSA CHIP cohort is comparable to other published cohorts.

Logistic regression models were constructed to analyse the impact of statin treatment on the likelihood of having high VAF (>=10%) versus low VAF (<10%) CHIP for *DNMT3A* and *TET2*, controlling for age, sex, serum cholesterol and cardiovascular risk factors and disease using data from waves 8 (2017) and 9 (2019), which represent the most complete and consistent timepoint in terms of sample availability, health outcome data and medication data. There were 494 and 555 individuals with *DNMT3A* and *TET2* mutated CHIP in this dataset, respectively, with demographic data provided in Figure 2A. The assumptions of linearity of independent variables and log odds of the dependent variable were met (Figure S1A-B) and analysis of generalised variance inflation factors (GVIFs) showed no significant co-linearity between included parameters in either model (Table S3). The odds ratios (ORs) of these gene specific multiparameter logistic regression models are shown in Figures 2B-C. Statin therapy was associated with a reduced risk of high VAF *TET2* CHIP in the multivariate model (OR (95% confidence interval (CI): 0.29 (0.15-0.56), p<0.001) and age-adjusted univariate model (p=0.013). Antiplatelet therapy was associated with an increased risk of high VAF *TET2* CHIP (OR (95% CI): 2.57 (1.36-4.90, p<0.01)) but this association was not robust in an age-adjusted univariate model (p=0.052). These associations were not observed in the *DNMT3A* logistic regression model.

**Figure 2.**
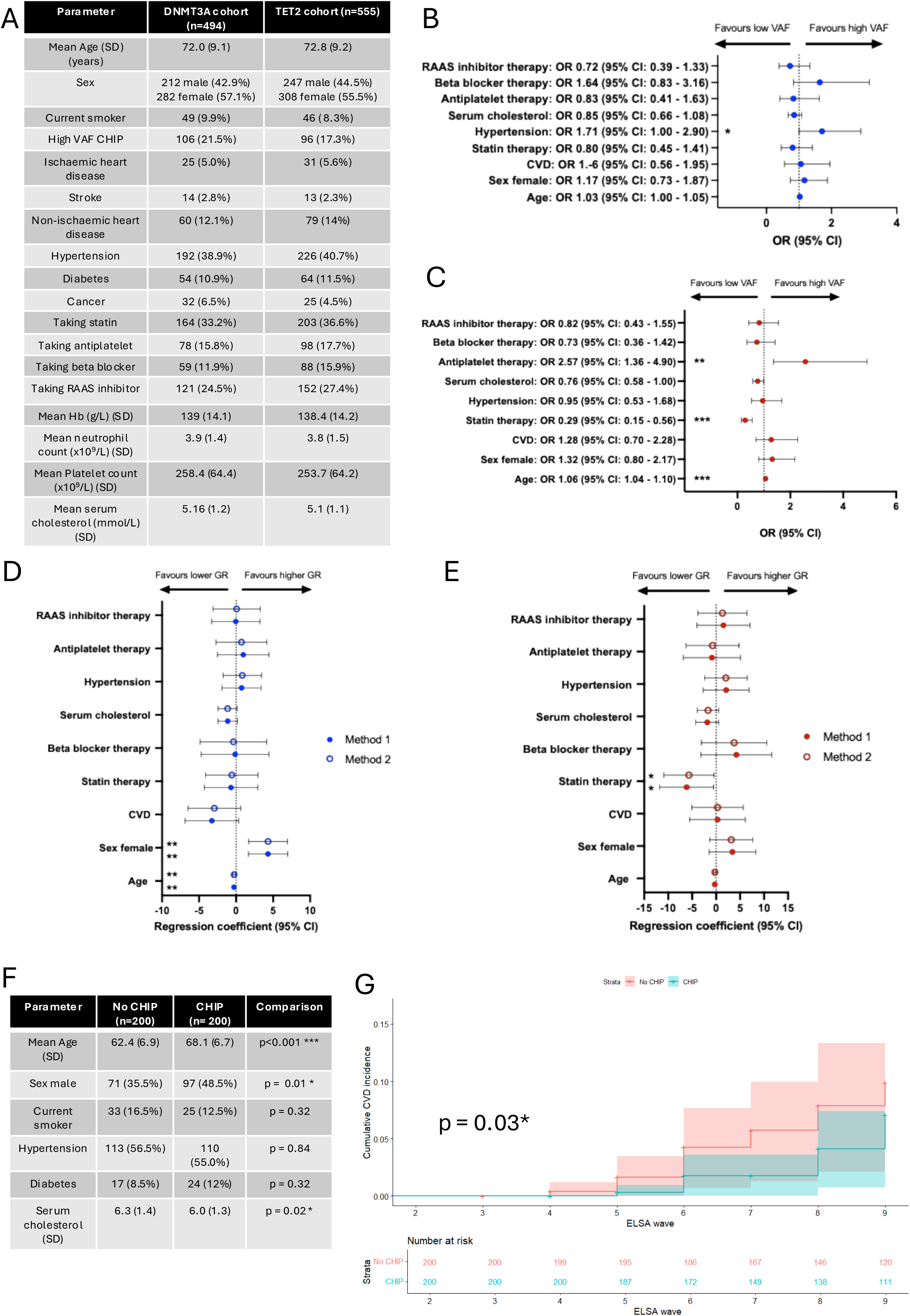
Analysis of the associations of statin therapy on CHIP clonal expansion and incident cardiovascular disease. A – Characteristics of DNMT3A and TET2 CHIP cohorts used in regression modelling of high VAF (>=10%) CHIP by gene. Ischaemic heart disease defined as a history of angina or myocardial infarction. Non-ischaemic heart disease defined as arrhythmias and valvular heart disease. RAAS: Renin-aldosterone-angiotensin system. B and C) Multivariate logistic regression model of high VAF vs low VAF DNMT3A (B) and TET2 CHIP (C) presented as odds ratios (OR) with 95% confidence intervals (CI). CVD defined as combined ischaemic heart disease, non-ischaemic heart disease and stroke. D and E) Robust multivariate regression coefficients of DNMT3A (D) and TET2 (E) CHIP clonal growth rate (GR) as calculated in Method 1 (described in Mack et at (2024)) and Method 2 (described by Uddin et al (2024)). F) Characteristics of cohort of ELSA wave 2 participants on statin primary prevention of CVD used to construct cox proportional hazards model, by CHIP status. Comparisons carried out with student’s t test for continuous variables and Chi-squared test for binary variables. SD: standard deviation. G) Cumulative incidence of CVD (defined as new diagnosis of myocardial infarction or stroke) in the cohort described in F, stratified by CHIP status and compared using the logrank test. *:p<0.05, **: p<0.01, ***: p<0.001.

To assess whether statin therapy is affects CHIP clonal growth rate we identified 120 and 93 individuals with *DNMT3A* and *TET2* CHIP and longitudinal sampling across successive ELSA waves respectively, and calculated CHIP clonal growth rate using two previously published methods(10),(7). Where mutations were undetectable at one timepoint, if the sequencing depth was >1000, the locus specific limit of detection (based on sequencing depth) was used as the VAF (Tables S5-6). Serial VAFs were corrected for the myeloid:lymphoid ratio in the full blood count parameters, taken as part of the same blood draw, to correct for any differences observed due to subclinical acute inflammatory responses. Model covariates included CVD comorbidities and commonly co-prescribed medications, with no evidence of co-linearity (Table S7). The mean follow-up time was 11.8 and 12.0 years for *DNMT3A* and *TET2*, respectively (Table S4). The mean growth rate for *TET2* was significantly higher than *DNMT3A* (9.1% and 2.9% per year, respectively, p<0.001, Table S4), in keeping with published data(4). Robust regression models were constructed to adjust for any other cardiovascular risk factors or commonly co-prescribed medication (Figures 2D-E). Similar to the logistic regression analysis, statin therapy was associated with a reduction in *TET2* clonal growth rate of 6.15% per year (p <0.05), with no significant association for *DNMT3A* CHIP growth rate. Age was minimally inversely associated with *DNMT3A* growth rate (regression coefficient (RC): -0.28, p<0.01) and female sex was associated with a higher *DNMT3A* clonal growth rate (RC:4.32, p<0.01), in line with the observation that *DNMT3A* mutant CHIP is more common in females(11).

To understand whether statin therapy had a differential benefit in clinical outcome data in individuals with CHIP, we constructed cumulative incidence curves of CVD (combined myocardial infarction and stroke) of 400 individuals in ELSA wave 2 who received statin therapy as primary prevention, stratified by CHIP status and matched using propensity score matching for age, sex, hypertension, diabetes, serum cholesterol and smoking status (Figure 2F). The mutational profile of individuals with CHIP was similar to the whole cohort (Figure S1 C-E). With 35 events observed across cases and controls, this cohort was not adequately powered to analyse gene-specific effects of statins. A cox proportional hazard model was constructed using age, sex and serum cholesterol as covariates, to correct for baseline differences. The cumulative incidence of CVD is shown in Figure 2G, indicating that statin primary prevention significantly reduces incident cardiovascular disease in individuals with CHIP compared to controls (p=0.03).

While similar analyses were not possible for HM due to the low incidence observed in the cohort, using data from these analyses in conjunction with the MN-predict tool(5) allows exploratory prediction of the potential myeloid malignancy relative risk reduction with statin treatment. Example myeloid malignancy risk predictions using the MN-predict “MDS demo” data and assuming a single *TET2* mutation with the mean growth rate observed in this study (9.1% per year), statin treatment for 10 years and a 6.15% reduction in clonal growth rate with statins are shown in Table S9. Allowing these assumptions, an individual with an initial *TET2* VAF of 10% could achieve a 33% relative risk of reduction of developing MDS.

In summary, in this study we sought to explore the impact of statins as extrinsic modifiers of CHIP clonal expansion. We characterise ELSA as a CHIP cohort with a characteristic CHIP mutational profile and epidemiological associations, which permits sequential mutation analysis in a long-term cohort of older participants, broadly selected from around England. We show that statin treatment is associated with a lower probability of having large *TET2* CHIP clones, and a lower rate of *TET2* CHIP clonal expansion in a gene specific manner. Whilst some other associations were also found (notably the relationship between antiplatelet therapy and high VAF *TET2* CHIP) that may warrant further investigation, these were less consistent and robust than the repeated association between *TET2* CHIP and statin therapy. These associations appear independent of serum cholesterol, which did not have any significant associations in these analyses, consistent with previous studies of determinants of clonal dynamics(7), raising the possibility that pleiotropic effects of statins could be contributing. Further, we show that in the ELSA cohort, primary statin therapy is associated with reduced incidence of cardiovascular disease in individuals with CHIP compared to controls, suggesting that statins may have a more specific action in CHIP than the general population. Extrapolation of the findings to myeloid malignant prediction tools suggests that statins could reduce the risk of malignant transformation in *TET2* CHIP. Whilst one cannot infer causality from associations and further mechanistic studies and validation in other cohorts and prospective randomised controlled trials is required, these analyses mirror epidemiological data suggesting that statins reduce the risk of malignant progression from MDS to AML(6), and represent a thought-provoking observation for the possibility of both CVD and malignant disease prevention in older adults with CHIP.

## Acknowledgements

The English Longitudinal Study of Ageing is funded by the National Institute on Aging (grant number RO1AG17644) and the National Institute for Health and Care Research (198/1074-02). EP is funded by the Cancer Research UK Advanced Clinician Scientist award A24873. ENM is funded by the Medical Research Council (grant reference MR/X001423/1). The authors thank the ELSA project team members, the National Centre for Social Research and ELSA participants for their time and commitment to the study. SD is supported by the BHF Data Science Centre / CVD-COVID-UK/COVID-IMPACT consortium, led by HDR UK (SP/19/3/34678), NIHR Biomedical Research Centre at University College London Hospital NHS Trust (UCLH BRC), BHF Accelerator Award (AA/18/6/24223), Multimorbidity Mechanism and Therapeutic Research Collaborative (MMTRC, grant number MR/V033867/1), NIHR-UKRI CONVALESCENCE study, and the Longitudinal Health and Wellbeing COVID-19 National Core Study, which was established by the UK Chief Scientific Officer in October, 2020, and funded by UKRI (grant references MC_PC_20030 and MC_PC_20059)

## Author contributions

Conceptualization: EN and EP; Methodology: EN, YH, PD, JH and EP; Investigation: EN, YH, PD; Writing – Original Draft, EN, Writing – Review & Editing: SD, AS, EP; Funding Acquisition, EN, AS and EP; Resources: AS; Supervision: EP.

## Data availability

The ELSA data is available on UK Data Service. All original code has been deposited at https://github.com/e-nuttallmusson/ELSA_statins_clonal_dynamics and is publicly available as of the date of publication. Any additional information required to reanalyse the data reported in this paper is available from the lead contact upon reasonable request, subject to General Data Protection Regulations.

