## supplementary material for "Analysis of the effects of statin therapy on clonal dynamics in clonal haematopoiesis of indeterminate potential: insights from the English Longitudinal Study of Ageing"

### SUPPLEMENTARY METHODS

### EXPERIMENTAL MODEL AND STUDY PARTICIPANT DETAILS

#### *ELSA samples*

ELSA participants were invited for peripheral blood (PB) sampling for genomic DNA extraction in waves 2 (2004), 4 (2008), 6 (2012), 8 (2017) and 9 (2019) of data collection. Ethical approval for each wave was approved by the South Central - Berkshire Research Ethics Committee and informed consent was obtained from all participants prior to each PB draw. Blood samples for genetic analysis were taken in waves 2, 4 and 6 (EDTA samples) and in waves 8 and 9 (Paxgene® samples). Whole PB samples were frozen at -80°C prior to DNA extraction. The age and sex demographics, and the sample numbers per wave are provided in figure 1A. Assays forming part of the ELSA dataset (including full blood count parameters, lipid profiles and markers of inflammation) were undertaken at the United Kingdom Accreditation Service approved Royal Victoria Infirmary laboratory in Newcastle, UK.

### METHOD DETAILS

#### DNA extraction and quantification

DNA was extracted from peripheral blood samples from waves 2, 4 and 6 (EDTA samples), and waves 8 and 9 (Paxgene® samples). DNA was extracted from samples in waves 2 and 4 by Source BioSciences. For samples from waves 6, 8 and 9, DNA was extracted from 200ul of whole blood using Agencourt® GenfindTM v2 kit on the Assist Plus Pipetting Robot (Integra Biosciences). All DNA samples were quantified using Invitrogen™ Quant-iT™ PicoGreen™ dsDNA Assay Kit and normalised to 50ng/ul using low TE buffer.

#### Myeloid NGS library preparation and sequencing

A custom myeloid amplicon library was designed by Fluidigm, including 641 amplicons covering 27 genes commonly mutated in myeloid malignancies and CHIP. Details of individual assays are included in Table S1. NGS libraries were constructed using the Fluidigm Juno Targeted DNA Sequencing Library Preparation System LP 192.24 IFCs. Libraries were cleaned up using Agencourt AMPure XP magnetic beads (Beckman Coulter PN A63880) and library quality was assessed with Agilent Bioanalyzer before sequencing on the NovaSeq 6000 Sequencing System (Novogene).

#### Variant calling

##### *Defining CHIP cases*

Fastq file quality was assessed with FastQC v0.11.8. Reads were aligned to a modified GRCh38 reference build with unchanged U2AF1 coordinates (1) using the bwa mem alignment algorithm (version 0.7.12-r1039). SAM files were converted to BAM files and then sorted and indexed using samtools (version 1.11). Coverage was assessed using both samtools depth and bedtools coverage (bedtools v2.30.0).

Variant calling was carried out using GATK (version 4.3.0.0) mutect2 according to the GATK Somatic short variant discovery (SNVs + Indels) best practices with the exception of marking duplicates (given the amplicon-based sequencing approach without unique molecular identifiers). The --max-reads-per-alignment-start argument was set at 0 given the amplicon sequencing approach. Files containing details of known SNPs and indels were downloaded from the relevant GATK resource bundle (Grch38/Hg38 Resources). Base (quality score) recalibration undertaken using known sites of SNPs and indels from the following files: Homo_sapiens_assembly38.dbsnp138.vcf, Homo_sapiens_assembly38.known_indels.vcf.gz, and Mills_and_1000G_gold_standard.indels.hg38.vcf.gz.

Calculation of the fraction of reads coming from cross-sample contamination was carried out with CalculateContamination. Variant filtering using FilterMutectCalls was then carried out undertaken without orientation filtering given the amplicon sequencing approach, which yields either all F1R2 or F2R1 reads at a given variant. Variants were filtered out if the coverage depth was less than 50.

Remaining variants then underwent additional annotation and filtering according to a published methods (2). Briefly, variant annotation was carried out using ANNOVAR (3) and the included cosmic70 database. Calls were filtered for presence on the 'whitelist' (2) (corresponding to a consensus on canonical CHIP driver mutations), using modified published R scripts to ensure those nonsense mutations designated as "stopgain" by mutect2 were not filtered out. Remaining calls were filtered out if they weren't supported on both the forward and reverse strands, had an alternate allele depth of < 20 or had a variant allele frequency < 0.02, in keeping with the consensus definition of CHIP. Further, variants were filtered out if they failed mutect2 'base quality' filter, if they failed the binomial test (suggesting a germline variant) or if they were known common artifacts (2). ASXL1 variants called at p.G645Vfs*58 and p.G646Wfs*12 (a common artifact) were filtered out if present at VAF of <0.1. Samples with ASXL1 exon 12 variants with identical read counts that were erroneously called at both p.G645Vfs*58 and p.G646Wfs*12 were considered to have one variant at p.G646Wfs*12. For multiallelic calls, the dominant VAF was considered and they were included if present in COSMIC (4). Variants flagged for manual review according to the whitelist filter were also included if they were present in COSMIC.

#### *Defining CHIP controls*

Controls were defined by the absence of variants identified using a similar filtering strategy as outlined above with some key differences. In order to ensure control samples didn't include potentially relevant variants that are not on the whitelist, this filter was not applied to the output of mutect2. Baseline quality filters were applied to remove likely sequencing artifacts (variants failing contamination, base_qual or weak_evidence filters, and those not supported on both the forward and reverse strands) and germline/benign variants (calls failing the germline filter and synonymous SNVs). Variants failing 'slippage' filter were filtered out if present at higher frequency than the common ASXL1 p.G646Wfs*12 with a VAF <0.05. Remaining SNV variants were assessed with Polyphen2 (5) to predict pathogenicity.

Samples were removed as candidate controls if they had a detectable whitelist variant or a variant specified for 'manual review' in the whitelist filtering strategy, an SNV not considered 'benign' by Polyphen2, any variant present in COSMIC and any variant present in the myelodysplastic syndrome cohort used to define the IPSS-M (6).

Remaining samples were considered as candidate controls if they had a mean sequencing depth across all amplicons of 800.

**Integrating variant calling and ELSA data**

ELSA survey and biomedical data were downloaded from the UK Data Service(7)(8). Comorbidities were defined to encompass whether people had an existing diagnosis, a previous diagnosis or were prescribed or advised medication to treat a condition. Additionally, conditions of a similar nature or pathophysiology were combined into summary variables (e.g. individuals with a history of a "heart attack" or a history of angina were designated as having ischaemic heart disease). Individuals were classed as smokers if they reported being current smokers.

Detailed medication data was available for waves 6, 8 and 9, coded according to the British National Formulary (BNF). Participants were considered to be taking a medication if it had been prescribed and taken in the week preceding the biomedical fieldworker visit.

CHIP cases and control samples were linked to the ELSA datasets through assigned unique ELSA identifiers. Repeat samples over successive ELSA waves were integrated longitudinally. Individuals with a diagnosis of a malignant haematological disorder were excluded from analyses of CHIP associations.

Assessment for association between the presence of CHIP and common comorbidities was assessed using wave 8 and 9 data and blood samples, which correspond to the largest quasi single time point. CHIP cases and controls were matched for age, sex and smoking status using nearest neighbour propensity score matching (matchIt R package). Odds ratios were calculated by conditional maximum likelihood estimation (Fisher) using the epitools R package. Plots were produced using GraphPad Prism.

### QUANTIFICATION AND STATISTICAL ANALYSIS

Analyses were performed using R 4.2.0 unless otherwise stated. Further information on regression and cox proportional hazard analyses performed are provided below.

**Regression analyses**

***Logistic regression analysis of determinants of high VAF CHIP cases***

CHIP cases identified in waves 8 and 9 were analysed for determinants of high VAF DNMT3A and TET2 CHIP cases. If an individual had more than one mutation in same gene, details of the dominant clone (highest VAF) was used in models. Where age>90 was censored by ELSA, age was conservatively assumed to be 90 years. Missing values for serum cholesterol measurements (4 and 5 individuals for the DNMT3A and TET2 cohort, respectively) were imputed with the median value. A multivariate logistic regression model was constructed around the binary dependent variable of VAF category, with 'high VAF' defined as a VAF >= 10% and 'low VAF' defined as a VAF < 10%. The number of covariates were limited by the frequency of high VAF cases in each cohort, with a minimum of 10 high VAF cases per covariate. Age, sex, cardiovascular disease, statin therapy, hypertension, serum cholesterol, antiplatelet therapy, beta blocker therapy and renin-angiotensin-aldosterone inhibitor therapy were included as independent variables. Included variables were assessed for collinearity using generalised variance inflation factors (GVIF) using gvif function in the 'glmtoolbox' package (v 0.1.11). Continuous predictor variables were plotted against the logit of the outcome to confirm the linearity of these relationships. Plots were produced using GraphPad Prism.

***Robust regression analysis of determinants of CHIP clonal growth rate***

CHIP cases identified in waves 8 and 9 were analysed for gene specific determinants of CHIP clonal growth rate. CHIP clonal growth rate was calculated for individuals with repeated blood sampling for individuals with the same mutation identified at multiple timepoints and individuals who were controls in early waves who subsequently acquired mutations (or vice versa), for whom comorbidity and medication data were available. Cases with splice variants were manually reviewed to ensure the same variant was detected over successive waves. Measured VAFs were corrected by the myeloid:lymphoid ratio from full blood count data taken at the same blood draw to reduce the effect of subclinical inflammatory responses on calculated clonal growth rate. Two published methods (described in Mack et at (2024)(9) and Uddin et al (2024)(10) were used to calculate growth rate as a sensitivity analysis. For those with more than one variant, the clone with the greatest clonal growth rate was considered. Those identified as controls at one timepoint were only included if the sequencing coverage at the locus at previous or subsequent mutations was greater than 1000 and therefore sensitive enough to detect variants at a VAF of 2%. Control samples meeting this criterion were conservatively assumed to have a VAF at the limit of detection of the assay, based on locus specific sequencing depth. Comorbidities were coded on most recent ELSA wave timepoint given the chronicity of the conditions and likelihood of relevant pre-diagnosis pathophysiology. Prescribed medication data were coded on wave 6 ELSA data to capture inter-timepoint information to reflect the effect of chronicity of exposure to medication. To ensure analyses were adequately powered, candidate categorical variables were included if >=10 observations were present in the dataset. Only cases with complete datasets were included. Included variables were assessed for collinearity using generalised variance inflation factors using gvif function in the 'glmtoolbox' package (v 0.1.11). Plots were produced using GraphPad Prism.

**Cox proportional hazard analysis of statin primary prevention on cardiovascular disease incidence by CHIP status**

Survey data from participants in ELSA wave 2 designated as CHIP cases and controls were integrated over ELSA waves 1-9. ELSA wave 2 was the first wave with DNA samples available. Individuals with a history of cardiovascular disease (defined as myocardial infarction or stroke) in ELSA waves 1 and 2 were excluded from analysis. Statin medication was recorded by wave based on BNF coded medications as previously described, and self-reported survey data (where available). Eligible individuals were considered to be taking statins as primary prevention if they were taking statins without a previous diagnosis of myocardial infarction or stroke. Eligible CHIP and controls were matched using propensity score matching for age, sex, hypertension, diabetes, serum cholesterol and smoking status. Any variables that remained significantly different were included in the cox proportional hazards model as covariates. The proportional hazards assumption was checked using the cox.zph function in the "survival" package (version

3.8-3). Individuals were censored if they were no longer under ELSA follow up. Cumulative incident curves were compared using the logrank test.

**FIGURE S1:**

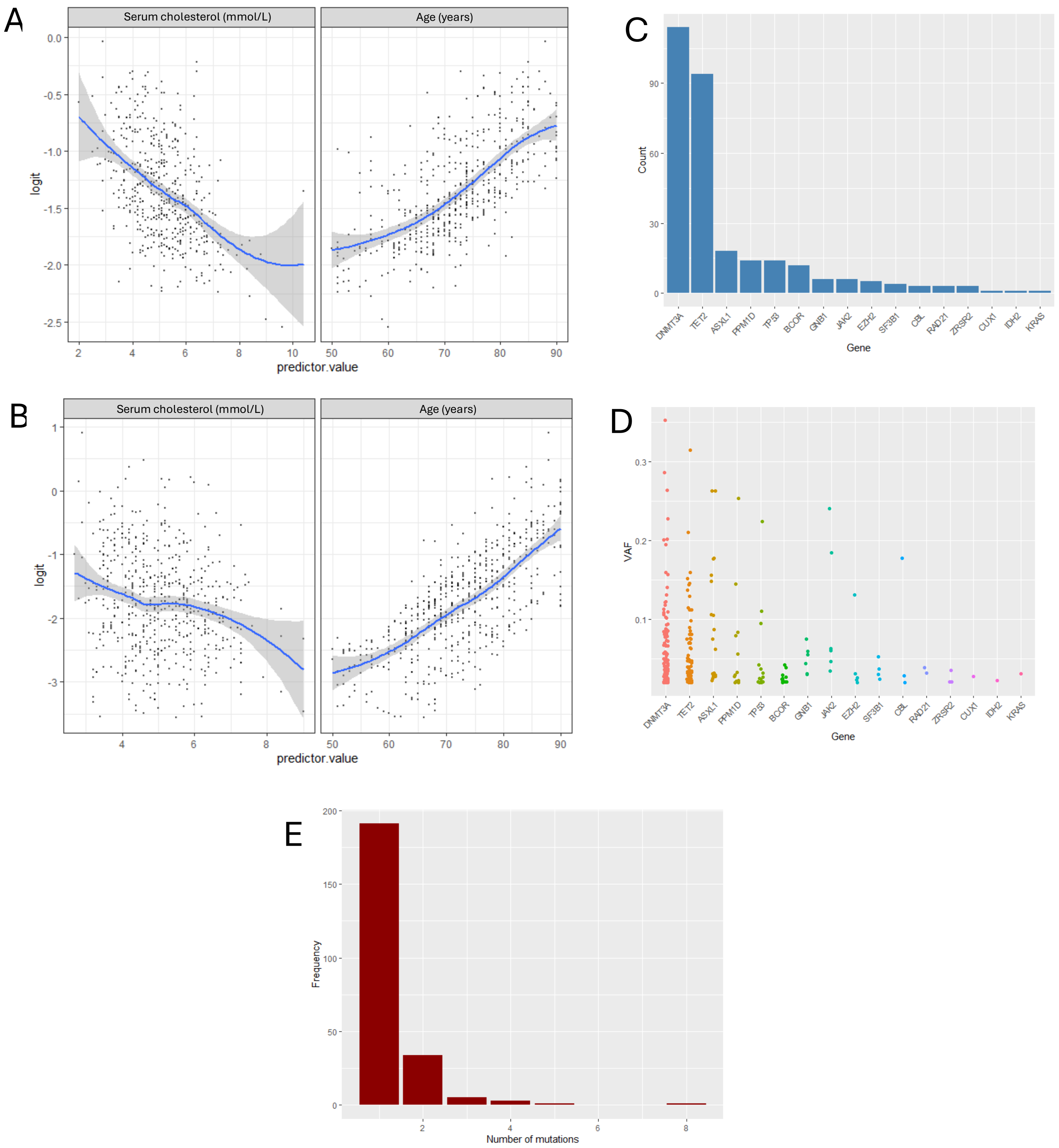


Figure S1:
A and B) Logits plotted against continuous variables included in multivariate logistic regression models of high VAF (>=10%) vs low VAF (<10%) CHIP illustrated in figure 2B-C for DNMT3A (A) and TET2 (B). C-E) Mutational profile of the cohort used for CVD (myocardial infarction and stroke) incidence analysis showing mutated gene frequency (C), VAF distributions (D) and mutation count per individual (E).

Table S3: Generalised variance inflation factors (GVIFs) of covariates in logistic regression models of high VAF vs low VAF CHIP for DNMT3A and TET2.

| Covariate | GVIF | df | GVIF^(1/(2*df)) |
|---|---|---|---|
| DNMT3A model | | | |
| Age | 1.135 | 1 | 1.0654 |
| Sex female | 1.1053 | 1 | 1.0513 |
| Cardiovascular disease | 1.2801 | 1 | 1.1314 |
| Statin therapy | 1.5242 | 1 | 1.2346 |
| Hypertension | 1.4615 | 1 | 1.2089 |
| Serum cholesterol | 1.6061 | 1 | 1.2673 |
| Antiplatelet therapy | 1.4083 | 1 | 1.1867 |
| Beta blocker therapy | 1.214 | 1 | 1.1018 |
| RAAS inhibitor therapy | 1.5519 | 1 | 1.2458 |
| TET2 model | | | |
| Age | 1.1579 | 1 | 1.0761 |
| Sex female | 1.1361 | 1 | 1.0659 |
| CVD | 1.1191 | 1 | 1.0579 |
| Statin therapy | 1.8229 | 1 | 1.3501 |
| Hypertension | 1.4884 | 1 | 1.22 |
| Serum cholesterol | 1.7152 | 1 | 1.3097 |
| Antiplatelet therapy | 1.4489 | 1 | 1.2037 |
| Beta blocker therapy | 1.1476 | 1 | 1.0713 |
| RAAS inhibitor therapy | 1.4716 | 1 | 1.2131 |

Table S4: Characteristics of cohorts with sequential sampling for growth rate calculation for DNMT3A and TET2 clones. Comparisons carried out with student's t test for continuous variables and Chi-squared test for binary variables. SD: standard deviation. Cardiovascular disease defined as combined ischaemic heart disease (angina and myocardial infarction), non-ischaemic heart disease and stroke. Mean growth rate calculated using Method 1 (described in Mack et at (2024)). ***: p<0.001.

| Parameter | DNMT3A (n=120) | TET2 (n=93) | Comparison |
|---|---|---|---|
| Mean Age (SD) (years) | 74.0 (7.0) | 75.6 (8.4) | p = 0.14 |
| Sex | Male: 49 (40.8%)<br>Female: 71 (59.2%) | Male: 38 (40.9%)<br>Female: 55 (59.1%) | p = 1.0 |
| Current smoker | 11 (9.2%) | 5 (5.3%) | p = 0.44 |
| Cardiovascular disease | 20 (16.7%) | 17 (18.3%) | p = 0.90 |
| Hypertension | 60 (50%) | 43 (46.2%) | p = 0.68 |
| Taking statin | 39 (32.5) | 36 (38.7%) | p = 0.43 |
| Taking antiplatelet | 23 (19.2%) | 21 (22.6%) | p = 0.66 |
| Taking beta blocker | 11 (9.2%) | 10 (10.7%) | p = 0.88 |
| Taking RAAS inhibitor | 33 (27.5%) | 28 (30.1%) | p = 0.79 |
| Mean Hb (g/L) (SD) | 139.4 (13.2) | 136.7 (13.4) | p = 0.14 |
| Mean neutrophil count ($x10^9$/L) (SD) | 4.0 (1.5) | 4.0 (1.6) | p = 0.33 |
| Mean Platelet count ($x10^9$/L) (SD) | 258.7 (65.2) | 249.5 (73.1) | p = 0.34 |
| Mean serum cholesterol (mmol/L) (SD) | 5.6 (1.2) | 5.4 (1.1) | p = 0.10 |
| Mean follow up (SD) (years) | 11.8 (2.7) | 12.0 (2.4) | p = 0.95 |
| Mean growth rate (% per year) (SD) | 2.9 (10.1) | 9.1 (13.7) | p < 0.001 *** |

Table S7: GVIFs of covariates in robust regression models of clonal growth rate for DNMT3A and TET2.

| Covariate | GVIF | df | GVIF^(1/(2*df)) |
|---|---|---|---|
| **DNMT3A model** | | | |
| Age | 1.3282 | 1 | 1.1525 |
| Sex | 1.291 | 1 | 1.1362 |
| CVD | 1.3709 | 1 | 1.1709 |
| Statin therapy | 2.1636 | 1 | 1.4709 |
| Beta blocker therapy | 1.303 | 1 | 1.1415 |
| Serum cholesterol | 1.9104 | 1 | 1.3822 |
| Hypertension | 1.3224 | 1 | 1.15 |
| Antiplatelet therapy | 1.4299 | 1 | 1.1958 |
| RAAS inhibitor therapy | 1.5796 | 1 | 1.2568 |
| Current smoker | 1.0427 | 1 | 1.0211 |
| **TET2 model** | | | |
| Age | 1.2646 | 1 | 1.1245 |
| Sex | 1.2904 | 1 | 1.136 |
| CVD | 1.1293 | 1 | 1.0627 |
| Statin therapy | 1.6912 | 1 | 1.3005 |
| Beta blocker therapy | 1.1818 | 1 | 1.0871 |
| Serum cholesterol | 1.5207 | 1 | 1.2332 |
| Hypertension | 1.2876 | 1 | 1.1347 |
| Antiplatelet therapy | 1.3982 | 1 | 1.1825 |
| RAAS inhibitor therapy | 1.4507 | 1 | 1.2045 |

Table S8: Confirmation that proportional hazards assumption met for Cox proportional hazard model of incidence CVD in individuals on statin primary prevention, stratified by CHIP status.

| Parameter | chisq | df | p |
|---|---|---|---|
| Age | 0.488 | 1 | 0.34 |
| Sex | 1.399 | 1 | 0.24 |
| Serum cholesterol | 0.108 | 1 | 0.73 |
| GLOBAL | 1.803 | 3 | 0.61 |

Table S9: MN-predict(11) results of extrapolated absolute risk (AR) and relative risk reduction (RRR) of myelodysplastic syndrome (MDS) and acute myeloid leukaemia (AML) for different TET2 VAFs

| **Starting TET2 VAF** | **No statin** | | | **Statin** | | | **RRR** | |
|---|---|---|---|---|---|---|---|---|
| | VAF after 10 years | 10 year MDS AR | 10 year AML AR | VAF after 10 years | 10 year MDS AR | 10 year AML AR | MDS | AML |
| 10% | 23.89172492 | 3% | 1% | 13.37406746 | 2% | 1% | 33% | 0% |
| 15% | 35.83758739 | 5% | 2% | 20.06110119 | 3% | 1% | 40% | 50% |
| 20% | 47.78344985 | 8% | 3% | 26.74813492 | 4% | 1% | 50% | 67% |

### SUPPLEMENTARY MATERIAL REFERENCES
